# The cardiac output – cerebral blood flow relation is abnormal in most ME/CFS patients with a normal heart rate and blood pressure response during a tilt test

**DOI:** 10.1101/2024.08.02.24311436

**Authors:** C. (Linda) M.C. van Campen, Freek W.A. Verheugt, Peter C. Rowe, Frans C. Visser

## Abstract

**Introduction:** Orthostatic intolerance is highly prevalent in patients with myalgic encephalomyelitis/chronic fatigue syndrome (ME/CFS) and is caused by an abnormal reduction in cerebral blood flow (CBF). In healthy controls (HC) regulation of CBF is complex and involves multiple mechanisms including cardiac output (CO), cerebral perfusion pressure, PO_2_ and PCO_2_, flow-metabolism coupling, and innervation of cerebral vessels. In ME/CFS multiple other mechanisms have also been identified.

**Aim of the study:** We previously found that both CBF and CO were reduced in ME/CFS patients during tilt testing, and we hypothesized that the relation between CBF and CO is abnormal and different from HC. In this retrospective study we analyzed this relation in a large group of patients. To compare the patient data with those of HC, we focused on patients with a normal heart rate (HR) and blood pressure (BP) response to upright tilt. Also, the influence of clinical data was analyzed.

**Methods:** A total of 534 ME/CFS patients and 49 HC underwent tilt testing with measurements of HR, BP, CBF, and CO. In 46 (9%) patients CO and CBF changes were in the normal range of HC, and in 488 (91%) an abnormal CO and CBF reduction was found.

**Results:** patients with a CO and CBF reduction in the range of HC had less severe disease and were more likely to be male. In patients with an abnormal CO and CBF reduction the slope of the regression line of CO versus CBF reduction was almost 1. A multiple regression analysis of the latter group, including patients with PetCO_2_ measurements (440/488: 90%) showed that the CO reduction for the major part predicted the CBF reduction, with a limited role for the PetCO_2_ reduction and the tilt duration. Other data did not add to the model.

**Conclusions:** Two different patient groups with a normal HR and BP response during the tilt were identified: those with a CO and CBF in the normal range of HC and those with an abnormal CO and CBF reduction during the tilt (91% of patients). The former group had milder disease and included more men. In the largest group of patients there was an almost 1:1 relation between the CO and CBF reduction, suggesting absence of compensatory vasodilation in the cerebral vasculature. This may be an appropriate target for clinical and therapeutic interventions.

## Introduction

Orthostatic intolerance is a well described phenomenon in patients with myalgic encephalomyelitis/chronic fatigue syndrome (ME/CFS) (Institute Of Medicine (IOM) 2015, van Campen, Verheugt et al. 2020, Wirth, Scheibenbogen et al. 2021). The symptoms of orthostatic intolerance—provoked by sitting, standing, upright activity, or tilt testing—are caused by an abnormal reduction in cerebral blood flow (CBF). The complex and only partially understood mechanisms of cerebral flow regulation have been studied in healthy controls and patients with a variety of diseases. Mechanisms involved include the cerebral perfusion pressure, PO_2_ and PCO_2_, flow-metabolism coupling, innervation of cerebral vessels, and blood viscosity (Hoiland, Fisher et al. 2019)(Hoiland et al. 2019). Also, age and gender influence CBF. In ME/CFS patients an abnormally decreased venous return due to the orthostatic stress (Timmers, Wieling et al. 2002, van Campen and Visser 2018), differences in blood volume (Streeten, Thomas et al. 2000, van Campen, Rowe et al. 2018), leg venous distensibility (Rowe, Barron et al. 1999, van Campen, Rowe et al. 2021), muscle blood pump characteristics (Keskin, Çiftçi et al. 2021), deconditioning (Fu, Vangundy et al. 2010), sympathetic drive (Wyller, Vitelli et al. 2016), the hemodynamic abnormality during a tilt test (van Campen, Verheugt et al. 2020), chronotropic incompetence (Davenport, Lehnen et al. 2019, van Campen, Verheugt et al. 2023), neuroinflammation (Nakatomi, Mizuno et al. 2014), autoimmunity of the nervous system (Wang, Ling et al. 2013), endothelial dysfunction (Scherbakov, Szklarski et al. 2020), disease severity (van Campen, Rowe et al. 2021), and microclots (Nunes, Kruger et al. 2022) may play a role.

Another important determinant of CBF is the cardiac output (CO). In a review Meng et al. described the relation between changes in CO versus changes in CBF in healthy controls (HC) during a variety of interventions, aiming to reduce or increase the CO (Meng, Hou et al. 2015). Summarizing the results of five studies with CO reductions in HC, the authors found that a reduction of 30% in CO resulted in a 10% reduction in CBF. On the other hand, Castle-Kirszbaum et al. found in their systematic review that “current literature is insufficiently robust to confirm an independent relationship between CO and CBF, and further studies with improved methodology are required before therapeutic interventions can be based on cardio-cerebral coupling” (Castle-Kirszbaum, Parkin et al. 2022).

In our previous studies we found that that the stroke volume index (SVI) and cardiac index (CI) reductions during a tilt test were larger in ME/CFS patients compared to HC (van Campen and Visser 2018, van Campen, Verheugt et al. 2023), confirming an earlier tilt test study in this population (Timmers, Wieling et al. 2002). We also studied the effect of orthostatic stress on CBF (van Campen, Rowe et al. 2020, van Campen, Rowe et al. 2020, van Campen, Verheugt et al. 2020, van Campen, Rowe et al. 2021, van Campen, Rowe et al. 2023). All studies showed that CBF was abnormally reduced during the orthostatic stress test in ME/CFS patients. Importantly, CI and CBF improved in these patients while wearing compression stockings, compared to the study without compression stockings (van Campen, Rowe et al. 2021), suggesting a causal role for the CO reduction on the CBF reduction. Based on these previous findings of a mean abnormal CBF and a mean abnormal CO reduction in ME/CFS patients, compared to HC, we hypothesized that there is a relation between the degree of CBF and the degree of CO changes -as part of the cerebral flow regulation (Meng, Hou et al. 2015, Claassen, Thijssen et al. 2021)-during a tilt test.

As HC studied in the present study have a normal heart rate (HR) and blood pressure (BP) response during the tilt test, we only analyzed the ME/CFS patients with a normal HR and BP response during the tilt and excluded those with postural orthostatic tachycardia syndrome (POTS), orthostatic hypotension (OH) or a (near)-syncope.

To measure CBF we used extracranial Doppler due to the inherent limitations of transcranial Doppler (Castle-Kirszbaum, Parkin et al. 2022). Moreover, apart from clinical data, HR, and BP, we also analyzed the influence of end tidal PCO_2_ (PetCO_2_), which has a powerful influence on CBF (Hoiland, Fisher et al. 2019).

## Patient, material, and methods

We reviewed the medical records of all ME/CFS patients who visited the out-patient Stichting CardioZorg clinic from October 2012 to May 2022 and who underwent a tilt-test. This clinic provides specialty assessment and treatment of individuals with ME/CFS. From the first visit, we determined whether participants satisfied the criteria for ME and for CFS (Fukuda, Straus et al. 1994, Carruthers, van de Sande et al. 2011), taking the exclusion criteria into account. No other illnesses were present that explained the symptomatology. Patients were selected for analysis when both Doppler data of CO and CBF were available both in the supine position and in the upright phase of the tilt test. No drugs influencing HR or BP were used at the time of the tilt testing. For comparison healthy controls were studied. Healthy controls were recruited from three sources: (a) announcements on ME/CFS patient advocacy websites, (b) posters in the medical clinic’s office building, and (c) healthy acquaintances of the ME/CFS participants. Subjects referred for syncope analysis or for other cardiologic diseases at our clinic were not considered as healthy controls. Disease severity in patients was scored according to the international consensus criteria (ICC), with severity scored as mild, moderate, severe, and very severe (Carruthers, van de Sande et al. 2011). Very severe patients (bedridden patients) were not studied here because they were not able to undergo a tilt test.

The study was carried out in accordance with the Declaration of Helsinki. All ME/CFS participants and healthy controls gave informed, written consent. The study was approved by the medical ethics committee of the Slotervaart Hospital, Amsterdam, for healthy controls P1450 and for ME/CFS patients P1736.

### Tilt-test protocol

Measurements were performed as described previously (van Campen, Verheugt et al. 2018, van Campen, Verheugt et al. 2020). Briefly, all participants were positioned for 20 min in a supine position before being tilted head-up to 70 degrees. Tilt duration was maximally 30 minutes, but in patients tilt duration was shorter and determined by the patient’s wellbeing and symptomatology; with increasing symptoms patients were tilted for a shorter period of time to avoid syncope and to avoid possible post-exertional malaise.

HR, systolic, and diastolic blood pressures (SBP, DBP) were continuously recorded by finger plethysmography (Eeftinck Schattenkerk, van Lieshout et al. 2009, Martina, Westerhof et al. 2012). After the test, HR and BP were extracted from the device and imported into an Excel spreadsheet. The changes in HR and BP during tilt testing were classified according to the consensus statement (Fedorowski, Burri et al. 2009, Freeman, Wieling et al. 2011, Sheldon, Grubb et al. 2015): normal HR and BP response (normal HR-BP response), classic orthostatic hypotension (cOH), delayed orthostatic hypotension (dOH), postural orthostatic tachycardia syndrome (POTS), and (near)-syncope. For comparison with HC, we only analyzed in the present study patients with a normal HR-BP response where blood pressures were within normal limits.

### Extracranial Doppler: cerebral blood flow measurements

Measurements were performed as described previously (van Campen, Verheugt et al. 2018, van Campen, Verheugt et al. 2020). Internal carotid artery and vertebral artery Doppler flow velocity frames were acquired by one operator (FCV), using a Vivid-I system (GE Healthcare, Hoevelaken, the Netherlands) equipped with a 6–13 MHz linear transducer. Frames were recorded in the supine position, before the onset of the tilt period, and while upright once or twice. When two sets of cerebral flow acquisitions were available, only the last set was analyzed. Blood flow of the internal carotid and vertebral arteries was calculated offline by one investigator (CMCvC). Blood flow in each vessel was calculated from the mean blood flow velocities times the vessel cross-sectional area and expressed in mL/minute. Flow in the individual arteries was calculated in 3-6 cardiac cycles and data were averaged. Total CBF was calculated by adding the flow of the four arteries.

### End-tidal CO_2_ pressure measurements

End-tidal CO_2_ pressures (in mmHg) were continuously measured with the Nonin Lifesense device. End-tilt pressures below 30 mmHg were considered abnormal (Stewart, Pianosi et al. 2018).

### Doppler echocardiographic measurements

Time velocity integral (VTI) frames were obtained in the resting supine position and at end of the tilt phase, immediately after the cerebral flow acquisitions. The aortic VTI was measured using a continuous wave Doppler pencil probe connected to a Vivid I machine (GE, Hoevelaken, NL) with the transducer positioned in the suprasternal notch. A maximal Doppler signal was assumed to be the optimal flow alignment. At least 2 frames of 6 seconds were obtained. Echo Doppler recordings were stored digitally. The VTI was measured off-line by manual tracing of at least 6 cardiac cycles, using the GE EchoPac post-processing software by one operator (CMCvC). The outflow tract diameter was manually drawn just below the valve insertion in the parasternal long-axis view of a previously made echocardiogram and the cross-sectional area calculated. As the outflow tract is not circular but ellipsoid, we used the data of Maes et al. (Maes, Pierard et al. 2017) to correct for the overestimation by the circular shape of the ellipsoid ventricular outflow tract calculation. In their study the overestimation of the outflow tract area, using the circular calculation by transthoracic echocardiography, was 24.5%. Therefore, we reduced the outflow tract area by 25%. Stroke volume (SV) was calculated from the aortic VTI, multiplied by the corrected aortic valve area, and expressed in mL. SV of the separate cycles were averaged. CO was calculated by the formula: SV times HR and expressed in L/min.

### Statistical analysis

Data were analyzed using the statistical package of IBM SPSS, version 29.0.00.0. All continuous data were tested for normal distribution using visual inspection of the Q-Q plots and presented as mean and standard deviation (SD) or as median with the interquartile range (IQR) where appropriate. Nominal data were compared using the Chi-square test (gender and disease severity, 3x2 and 3x3 tables). Group differences were explored using Welch ANOVA, by the Mann-Whitney U test in case of the comparison of two groups or by the Kruskal-Wallis test in the comparison of three or more groups. Post-hoc tests were performed using the Tukey or Dunn test. Multiple regression analysis was performed according to the guidelines of Laerd Statistics (Laerd Statistics 2015). Due to the large number of comparisons, to reduce type I errors, we choose a conservative p-value of <0.01 to be statistically significant.

## Results

From our database we identified 1135 ME/CFS patients who visited our clinic between October 2012 and May 2022, who fulfilled the criteria for both ME and CFS (Fukuda, Straus et al. 1994, Carruthers, van de Sande et al. 2011), and who had a tilt test because of the suspicion of orthostatic intolerance. In all patients CBF measurements as well as suprasternal derived SV were available. From this group we selected patients with a normal HR-BP response (n=664). Patients younger than 18 years were excluded leaving 629 patients. Also, patients with a BMI> 40 were excluded, leaving 612 patients. Patients with insufficient quality of the CBF or SV measurements were excluded as well as patients with missing data of CBF and SV, leaving 585 patients. Finally, 51 patients using HR and BP lowering drugs or lung medication containing sympathomimetics were excluded. This left 534 patients to be analyzed. For comparison with the patients 49 healthy controls (HC) with a normal HR-BP response, and with a complete set of CO and CBF measurements were also analyzed.

Based on the distribution of the %CO reduction versus the %CBF reduction (see Figure 1) we decided to separate patients in a group where the %CBF reduction was in the normal range of healthy controls, and a group of patients with a %CBF reduction below the lower limit of normal of healthy controls (cut-off value of %CBF reduction of -15%). Table 1 shows the baseline characteristics of the three groups. Four hundred and eighty-eight patients had a larger than normal %CBF reduction, while 46 showed a %CBF reduction in the normal range of HC. There were significantly more males in the patient group with a %CBF reduction in the normal range. Patients with an abnormal %CBF reduction were more severely affected by the disease, with a higher percentage of moderate and severe disease than patients with a %CBF reduction in the normal range. All other baseline characteristics were similar between the three groups.

**Figure 1:**
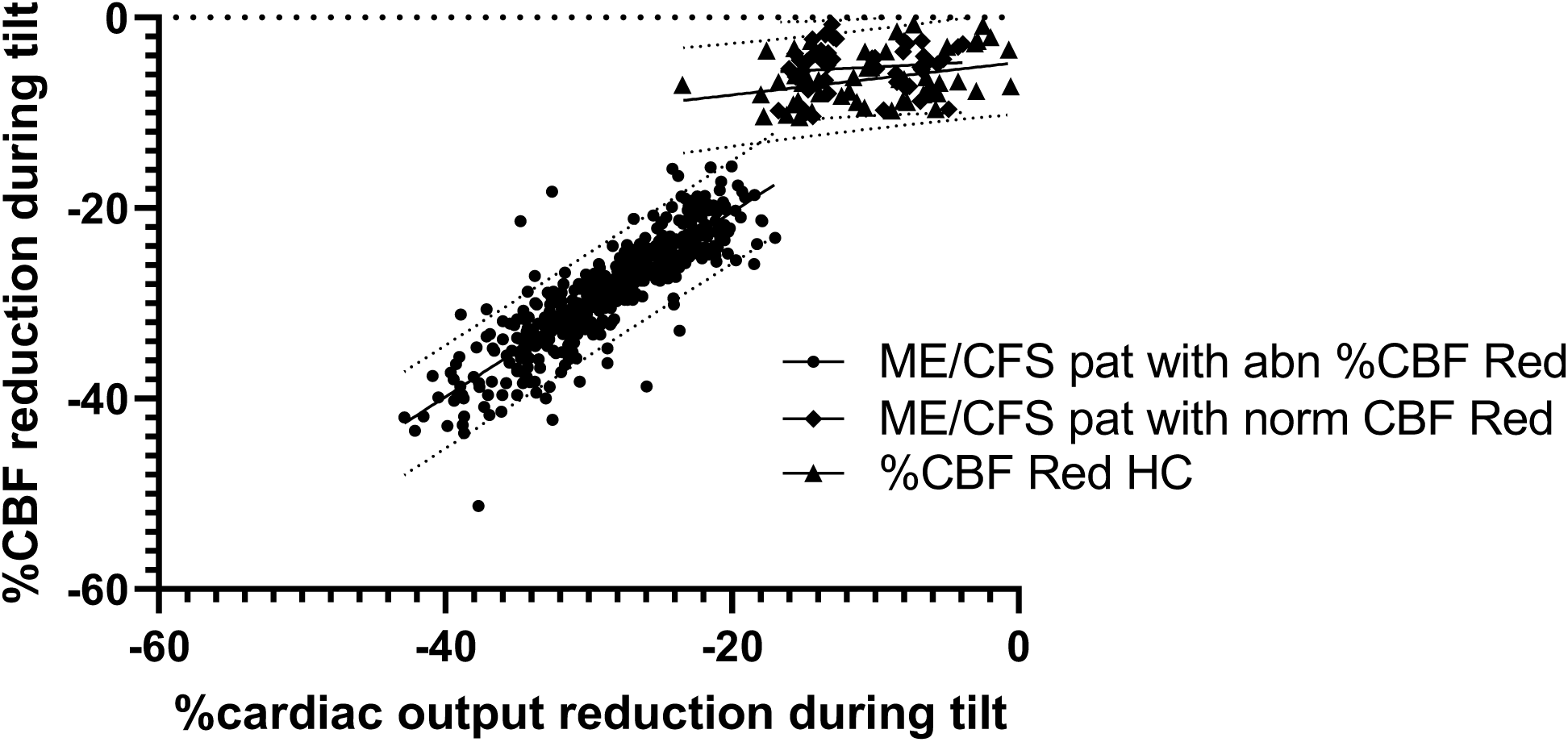
Percent cerebral blood flow reduction vs percent cardiac output reduction during the tilt test in ME/CFS patients and healthy controls. abn: abnormal; %CBF Red: percent reduction in cerebral blood flow during the tilt; HC: healthy controls; pat: patients; ME/CFS: myalgic encephalomyelitis/chronic fatigue syndrome; norm: normal.

**Table 1:** Baseline characteristics of ME/CFS patients with a %CBF reduction in the normal range of healthy controls during a tilt test, patients with an abnormal %CBF reduction, and healthy controls.

|  | ME/CFS with abnormal %CBF reduction (n=488) | ME/CFS with normal %CBF reduction (n=46) | Healthy controls (n=49) | p-value: Chi-square/<br>One-way Welch ANOVA with post-hoc; Kruskal-Wallis with post-hoc |
| --- | --- | --- | --- | --- |
| Male/female * | 70/418 (14/86%) | 19/27 (41/59%) | 11/38 (22/78%) | p<0.001 |
| Age (years) ∞ | 41 (12) | 44 (11) | 39 (15) | F (2, 580) = 1.540 p=0.215 |
| Height (cm) ∞ | 171 (8) | 174 (10) | 173 (7) | F (2, 580) = 3.377 p=0.035 |
| Weight (kg) # | 70 (62-81) | 74 (62-85) | 73 (63-82) | X<sup>2</sup> (2) = 1.788 p=0.409 |
| BMI (kg/m<sup>2</sup>) # | 23.8 (21.1-27.7) | 24.2 (21.2-27.4) | 24.3 (21.3-27.6) | X<sup>2</sup> (2) = 0.089 p=0.956 |
| BSA (m<sup>2</sup>) ∞ | 1.84 (0.20) | 1.88 (0.21) | 1.87 (0.19) | F (2, 580) = 1.592 p=0.204 |
| Disease duration (years) # | 12 (6-20) | 13 (7-21) | - | p=0.76 |
| Disease severity ©* | 141/263/84<br>(29/54/17%) | 35/10/1<br>(76/22/2%) | - | p<0.001 |

Footer table 1: \* Chi-square 2x2 or 2x3 analysis; ∞: One-way Welch ANOVA with Tukey/Games-Howell
post-hoc; #: Median (IQR) Mann-Whitney U or Kruskal-Wallis test with Dunn post-hoc; BMI: body mass
index; BSA: body surface area (formula duBois); CBF: cerebral blood flow; ME/CFS: myalgic
encephalomyelitis/chronic fatigue syndrome; ©: disease severity presented as the % with
mild/moderate/severe disease according to the ICC criteria (Carruthers, van de Sande et al. 2011). A
p-value of <0.01 is considered statistically significant.

Table 2 shows the hemodynamic data of the three groups in the supine position and at the end of the tilt phase. Supine and end-tilt HR were significantly higher in the patients with an abnormal %CBF reduction than in HC. Supine CO was higher in patients with an abnormal %CBF reduction during the tilt compared to HC. End-tilt CO was lowest in patients with an abnormal %CBF reduction compared to patients with a normal %CBF reduction and compared to HC. Accordingly, the %CO reduction at end-tilt was significantly larger in the patient group with an abnormal %CBF reduction. By definition, the end-tilt CBF was lower and the %CBF reduction larger in the patient group with the abnormal %CBF reduction compared to the other patient group and HC. The PetCO_2_ reduction was significantly larger in patients with an abnormal %CBF reduction than those with a %CBF reduction in the normal range of HC and compared to the PetCO_2_ reduction of HC. Also, the %patients with an PetCO2 <30 mmHg was larger in patients with an abnormal %CBF reduction. End-tilt DBP were higher in both patient groups compared to HC but reached significance only in the patient group with an abnormal %CBF reduction. End-tilt MAP was significantly higher in both patient groups compared to HC, but only reached significance in patients with an abnormal %CBF reduction. Tilt duration was significantly shorter in patients with an abnormal %CBF reduction compared to patients with a %CBF reduction in the normal range and compared to HC. Frame acquisition for CBF measurements lasted 3.3 (1.6) minutes without differences between groups (data not shown). Acquisition of VTI measurements lasted 0.7 (0.3) minutes without differences between groups (data not shown).

**Table 2:**
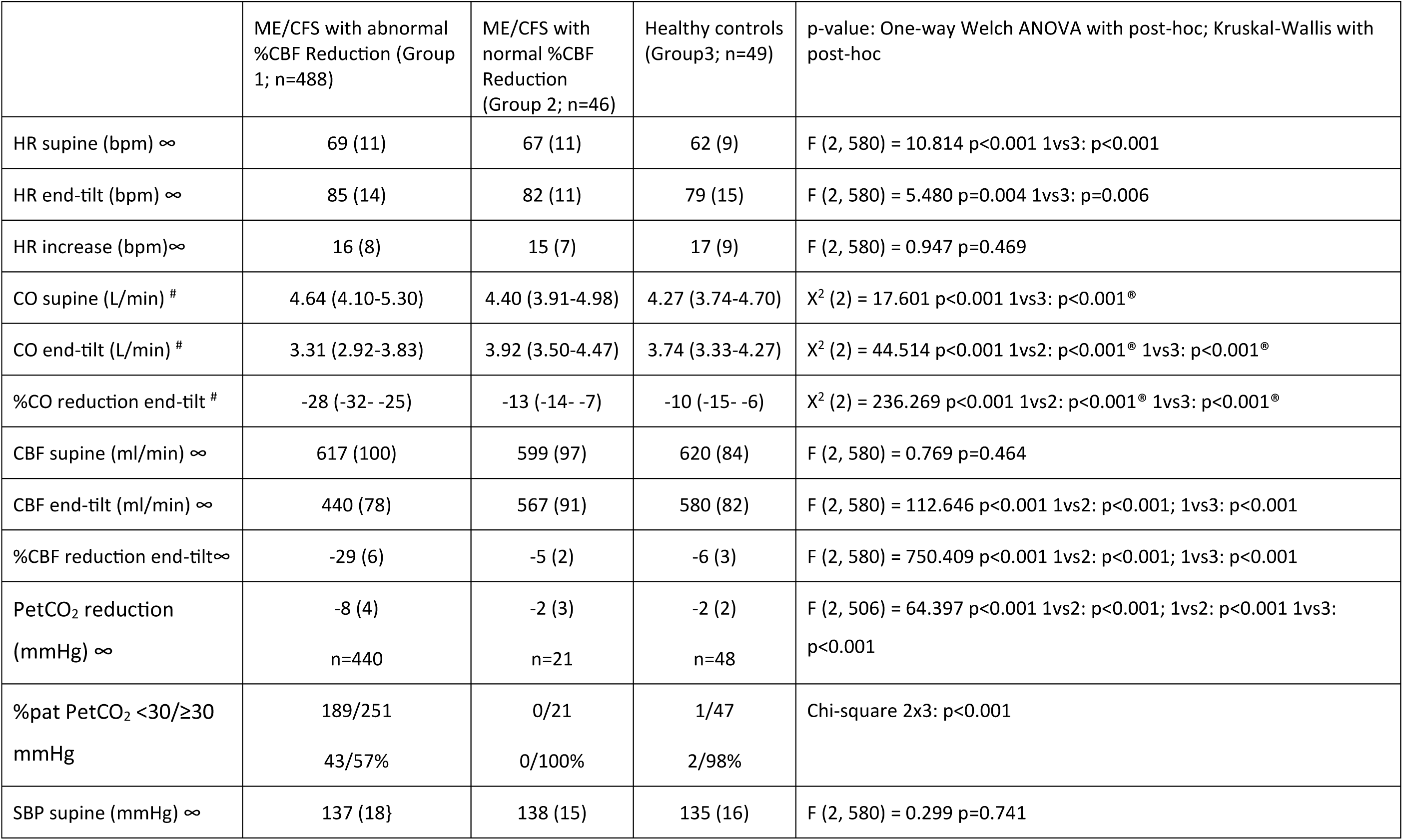
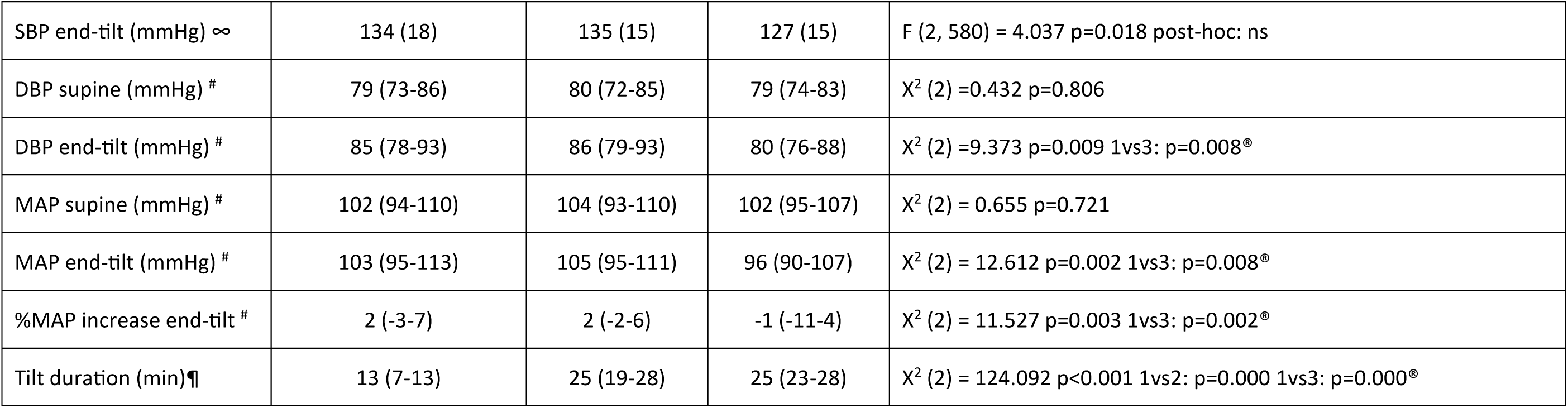
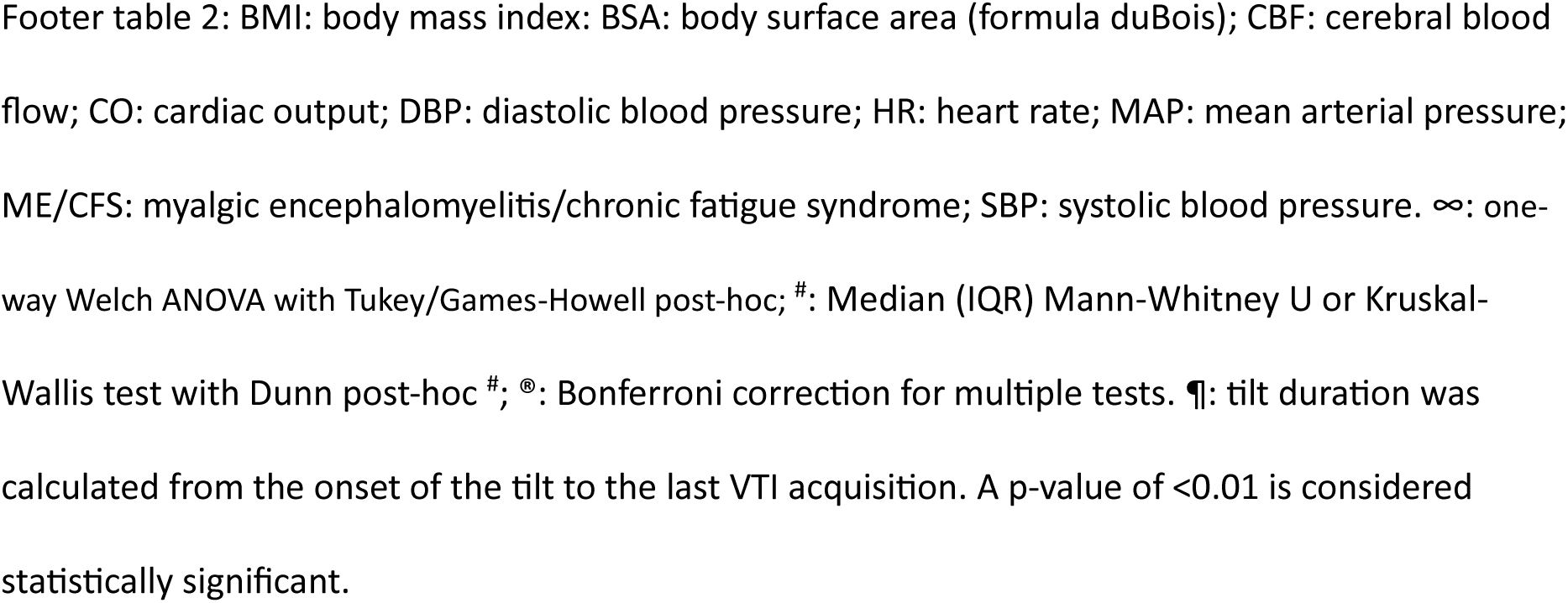
Tilt test data of ME/CFS patients with a %CBF reduction in the normal range of healthy controls during a tilt test, patients with an abnormal %CBF reduction, and healthy controls.

Figure 1 shows the relation between the %CO reduction and %CBF reduction in the three groups. In patients with an abnormal %CBF reduction the relation was highly significant: %CBF reduction = 0.971*%CO reduction – 1.027; R^2^ = 0.762; p<0.001. In patients with a normal %CBF reduction the slope was not significantly different from zero: %CBF reduction = 0.076*%CO reduction – 4.420; R^2^ = 0.014; p=0.438. In HC the slope was marginally significant: %CBF reduction = 0.171*%CO reduction – 4.708; R^2^ = 0.114; p=0.018.

To determine whether clinical and other hemodynamic variables were associated with the %CBF reduction during the tilt we analyzed the relation between %CBF reduction in the patient group with an abnormal %CBF reduction. Patients with a normal %CBF reduction were not analyzed as the regression line was not different from zero. In this subgroup of patients with an abnormal %CBF reduction 440/488 (90%) had adequate PetCO_2_ measurements (43% of these 440 patients had a PetCO_2_ below 30 mmHg). The %CBF reduction was related to gender, age, disease duration and severity, length, weight, the heart rate increase, the %CO reduction, PetCO_2_ reduction, patients with a PetCO2 < 30 mmHg, the %MAP increase, and tilt duration. Table 3 shows the results of the univariate analysis: increasing disease severity was associated with a larger %CBF reduction, a larger %CO reduction, a larger PetCO_2_ reduction, and an end-tilt PetCO_2_ < 30 mmHg resulted in significantly larger %CBF reductions. Other variables showed no significant relations. For the multivariate analysis variables were included that showed a p value < 0.1 in the univariate analysis. Table 4 showed that the %CO reduction, and tilt duration significantly predicted the %CBF reduction. The PetCO2 reduction showed a marginal contribution (p=0.011). All other variables did not significantly contribute to the model.

**Table 3:** Analysis of the possible relation between percent cerebral blood flow reduction during the tilt test and clinical and hemodynamic variables in ME/CFS patients with a normal heart rate and blood pressure response during the tilt and an abnormal CBF reduction.

| Variable | N=440<sup>®</sup> |  |  |
| --- | --- | --- | --- |
| Gender: M | %CBF=-0.306*F -28.762 | R<sup>2</sup>=0.000 | p=0.686 |
| Age (years) | %CBF=0.042*age -30.536 | R<sup>2</sup>=0.008 | p=0.054 |
| Disease duration (years) | %CBF=0.008*DisDur -28.697 | R<sup>2</sup>=0.000 | p=0.756 |
| Disease severity: mild © | %CBF=-1.516*moderate -2.537*severe –<br>27.492 | R<sup>2</sup>=0.024 | p=0.005 |
| Length (cm) | %CBF=-0.004*length -28.109 | R<sup>2</sup>=0.000 | p=0.897 |
| Weight (kg) | %CBF=0.022*weight -30.380 | R<sup>2</sup>=0.004 | p=0.188 |
| HR increase (bpm) | %CBF=-0.051*HR increase -29.606 | R<sup>2</sup>=0.006 | P=0.12 |
| %CO reduction | %CBF=0.974*%CO reduction -0.908 | R<sup>2</sup>=0.778 | p<0.001 |
| PetCO<sub>2</sub> reduction (mmHg) | %CBF=0.324*PetCO<sub>2</sub> reduction -26.328 | R<sup>2</sup>=0.062 | p<0.001 |
| End-tilt PCO<sub>2</sub> < 30 mmHg | %CBF=-2.361*PetCO<sub>2</sub>E-T – 27.792 | R<sup>2</sup>=0.045 | p<0.001 |
| %MAP increase (mmHg) | %CBF=-0.036*%MAP increase -28.709 | R<sup>2</sup>=0.003 | p=0.250 |
| Tilt duration (min) ¶ | %CBF=0.092*TiltDur -30.068 | R<sup>2</sup>=0.012 | p=0.020 |

Footer table 3: %CBF: percent cerebral blood flow reduction during tilt compared to supine CBF; CO: cardiac output; DisDur: disease duration; F: female; HR: heart rate; M: male; MAP: mean arterial pressure; ME/CFS: myalgic encephalomyelitis/chronic fatigue syndrome; PetCO<sub>2</sub>: end-tidal CO<sub>2</sub> pressure; PetCO<sub>2</sub>E-T: end-tilt PetCO<sub>2</sub> SBP: systolic blood pressure; TiltDur: tilt duration; ©: Disease severity according to the ICC criteria (Carruthers, van de Sande et al. 2011), ¶: tilt duration was calculated from the onset of the tilt to the last VTI acquisition, <sup>®</sup>: subset of patients with a complete PetCO<sub>2</sub> set (90% of all patients).

**Table 4:** Multiple logistic regression in ME/CFS patients with an abnormal percent cerebral blood flow reduction to predict the percent cerebral blood flow reduction during the tilt.

|  | B | t | p value |
| --- | --- | --- | --- |
| (Constant) | -0.048 | -0.048 | p=0.962 |
| Age (years) | 0.015 | 1.437 | p=0.151 |
| Dummy Moderate * | -0.307 | -1.003 | p=0.316 |
| Dummy Severe * | -0.663 | -1.731 | p=0.084 |
| %CO reduction | 0.959 | 37.399 | p<0.001 |
| PetCO<sub>2</sub> reduction (mmHg) | 0.100 | 2.555 | p=0.011 |
| End-tilt PetCO<sub>2</sub> <30 mmHg | -0.083 | -0.265 | p=0.791 |
| Tilt duration (min) ¶ | -0.063 | -3.008 | p=0.003 |

Footer table 4: %CO: percent cardiac output reduction during the tilt compared to supine; ME/CFS: myalgic encephalomyelitis/chronic fatigue syndrome; PetCO<sub>2</sub>: end-tidal CO<sub>2</sub> pressure. \* Disease severity according to the ICC criteria (Carruthers, van de Sande et al. 2011), ¶: tilt duration was calculated from the onset of the tilt to the last VTI acquisition.

## Discussion

We have demonstrated in different studies in ME/CFS patients that 1) there is a significantly larger reduction in cardiac index during a tilt test compared to HC (van Campen, Verheugt et al. 2023) (van Campen and Visser 2018), and 2) there is a significantly larger reduction in CBF during the tilt test compared to HC (van Campen, Verheugt et al. 2020, van Campen, Rowe et al. 2021, van Campen, Rowe et al. 2023). In the present study the CO and CBF data were compared between the patients and HC. Therefore, we selected those patients and HC that showed a normal HR-BP response.

Meng et al. have shown in their review of HC that a 30% CO reduction leads to an approoximately 10% reduction in CBF (Meng, Hou et al. 2015). Theoretically, the larger CBF reduction in ME/CFS patients follows the larger CO reduction, like the changes as in HC (30%CO reduction/10%CBF reduction), or, alternatively, a larger than expected CBF reduction is present. In a previous study the %CBF reduction was -23.6% in ME/CFS patients with a normal HR-BP response during a tilt test (van Campen, Verheugt et al. 2020). Therefore, we hypothesized that ME/CFS patients with a normal HR-BP response would show a larger %CBF reduction than expected by the %CO reduction.

In the present study two different patterns in patients were observed: first, only 9% of ME/CFS patients had %CBF reduction (-5%) and a % CO reduction (-13%) that fell within the range of HC, _w_ithout significant differences between these patients and HC (Table 2). In contrast, 91% of patients showed a large and abnormal %CBF reduction during the tilt, with a mean %CBF reduction of -29% and a %CO reduction of -28%.

The data of HC in the present study were compared with those in the review by Meng (Meng, Hou et al. 2015). The %CBF reduction in the five reviewed studies varied between -4 and -19%, the %CO reduction between -18 and -44, while in our study the %CBF reduction was -6% and the %CO -10%. However, there are differences in techniques used for both the CBF (transcranial Doppler vs extracranial Doppler in our study) and the CO measurements (inert gas/acetylene rebreathing, finger plethysmography, and impedance cardiography vs suprasternal Doppler in our study). Also, the orthostatic stressor was different between our study (tilt test) and the previous ones (mainly lower body negative pressure (LBNP) and standing up). These differences in measurement techniques and stressors may account for the differences in obtained CO and CBF data. For example, in a direct comparison study between LBNP and a tilt test, the CBF reduction by LBNP in HC was significantly less than during tilt testing, which the authors attributed to differences in PetCO_2_ (Bronzwaer, Verbree et al. 2017). Also, we demonstrated that CO by finger plethysmography underestimates the CO changes during the tilt compared to suprasternal Doppler (van Campen, Verheugt et al. 2021).

When comparing ME/CFS patients with a %CBF reduction in the normal range of HC and patients with an abnormal %CBF reduction, patients with a %CBF reduction in the normal range of HC had less severe ME/CFS and were more likely to be male than patients with an abnormal CBF and CO reduction. We have recently shown that disease severity, classified according to the ICC criteria (Carruthers, van de Sande et al. 2011), was related to the %CBF reduction: severe patients had a larger %CBF reduction during the tilt test than patients with a mild or moderate severity of the disease (van Campen, Rowe et al. 2023). The association between increasing symptom severity and a larger %CBF reduction is strengthened by this study showing that patients with worsening symptoms over time (including those with initially a %CBF reduction in the normal range) had a larger %CBF reduction during the second tilt test (van Campen, Rowe et al. 2023). Furthermore, Streeten et al. and our group showed that the use of military anti-shock trousers/compression stockings improved both OI and ME/CFS symptomatology and CBF abnormalities in ME/CFS patients (Streeten, Thomas et al. 2000, van Campen, Verheugt et al. 2018, van Campen, Rowe et al. 2021).

Although there is an association between ME/CFS disease severity and the %CBF reduction, there is no evidence that there is a cause-effect relation. This is evident from this patient group without an abnormal CBF reduction, but with symptoms varying between mild and severe disease.

The observation that males have less severe ME/CFS has been observed previously, reflected by less symptoms and a higher physical functioning scale than women (Buchwald, Pearlman et al. 1994, Tseng and Natelson 2004, Faro, Sàez-Francás et al. 2016). The mechanism of milder symptomatology in men is unknown, as is the observation of a lower prevalence of the disease in men.

The largest group of ME/CFS patients are those with an abnormal %CBF reduction during the tilt. As shown in Figure 1 the %CBF reduction parallels the %CO reduction with a slope between the %CBF and %CO reduction that is near 1. In the subgroup analysis of patients in whom also PetCO2 data were available (n=440) clinical variables like disease severity and an abnormal PetCO2 (<30 mmHg) were associated with the abnormal %CBF reduction (see table 3). However, the multiple regression analysis showed that only the %CO reduction was significantly associated with the %CBF reduction. The other variables did not contribute significantly.

One may divide mechanisms that regulate CBF into four distinct components or adaptive responses: autoregulation, chemoregulation (also called vascular reactivity), neuronal regulation (including the neurovascascular coupling and the effects of autonomic and sensory nerves on the extraparenchymal segments of the cerebral vasculature), and endothelium-dependent regulation (Claassen, Thijssen et al. 2021, Silverman and Petersen 2024). Autoregulation refers to how CBF responds to changes in blood pressure. In the present study there are minimal changes in DBP, DBP, and MAP, suggesting that autoregulation plays a minor role here. Second, the chemoregulation by CO_2_ (vascular reactivity) is probably normal in these patients as in the univariate analysis the %CBF reduction was related to the PetCO_2_ reduction and the mean PetCO_2_ reduction was 8 mmHg and the %CBF reduction was -29%. This relationship is in line with a number of studies in HC, where a 1 mmHg reduction in PetCO_2_ was related to a 3-4% reduction in CBF (see review Hoiland et al. (Hoiland, Fisher et al. 2019). Nevertheless, in the multivariate analysis the PetCO_2_ reduction was marginally related to the %CBF reduction due to our choice to set the significance level to 0.01, to reduce false positive findings. Third, the neurovascular coupling describes the close temporal and regional linkage between neural activity and CBF responses, where an increase in neuronal activity leads to an increase in CBF by dilatation of upstream pial arteries and near-by arterioles and capillaries (Willie, Tzeng et al. 2014, Phillips, Chan et al. 2016). In humans the neurovascular coupling can be studied e.g. using a visual stimulation with an eye-open and eye-closed protocol. Although in our protocol leg muscle tone may increase during standing, thereby increasing regional CBF, the large reduction in CO and CO_2_, together with the assessment of global CBF prevents assessment of neurovascular coupling. Fourth, the cerebrovascular endothelium exerts a profound influence on cerebral vessels and cerebral blood flow via smooth muscle cell dilators and constrictors, like NO, and endothelin-1 (Andresen, Shafi et al. 2006, Bai, Yu et al. 2022). Even under conditions of high orthostatic stress cerebral flow regulation in HC is preserved, as a study of Brown et al. showed that a mean %CO reduction of -44% resulted in a %CBF reduction of -19% (Brown, Dütsch et al. 2003). Giving the observation that the %CBF reduction is almost 1:1 related with the %CO reduction in over 90% of the ME/CFS patients with a normal HR-BP response, it is most likely that endothelial dysfunction of the cerebral vasculature plays a dominant role in the abnormal cerebral flow regulation during orthostatic stress. As outlined in the introduction, the mechanisms of cerebral flow regulation in HC are complex, but in ME/CFS patients additional factors that may disturb cerebral flow regulation, like abnormal venous return, blood volume changes, venous distensibility, deconditioning, chronotropic incompetence, neuroinflammation, autoimmunity, and microclots, can also play a role. Endothelial dysfunction in the brain has been described in a variety of cerebral diseases like Alzheimer disease (Fang, Hsieh et al. 2023), cerebral small vessel disease (Bai, Yu et al. 2022), in multiple sclerosis (Dubchenko, Ivanov et al. 2020), and possibly in Parkinson disease (Kollár, Blaho et al. 2022). In ME/CFS patients a number of studies have shown endothelial dysfunction using flow-mediated vasodilation/post-occlusive hyperemia (Newton, Kennedy et al. 2012, Scherbakov, Szklarski et al. 2020, Sørland, Sandvik et al. 2021, McLaughlin, Sanal-Hayes et al. 2023, Sandvik, Sørland et al. 2023). Our study, specifically targeting the cerebral vasculature, adds to evidence that endothelial dysfunction of the brain may be present in the majority of patients.

Figure 1 shows that there is a separation between patients with and without an abnormal %CBF reduction with a cut-off value of the %CBF reduction around -15%. This observation is strengthened by our previous study where patients with worsening symptoms showed a substantial further reduction in %CBF (van Campen, Rowe et al. 2023). Twenty-five of these patients with worsening symptoms (n= 71) initially had a %CBF reduction in the normal range of HC. In this subgroup the initial %CBF reduction was -6% (SD 4%) but changed to -25% (SD 5%), a value well beyond the cut-off value of -15%. Possibly one of the above-mentioned mechanisms in ME/CFS may become operational (venous return, hypovolemia, deconditioning, sympathetic activation, chronotropic incompetence, neuro-inflammation, auto-immunity, endothelial dysfunction, and micro clots), but this needs to be studied further.

Finally, the multiple regression shows that the %CBF change was inversely correlated with the tilt duration. Although the slope of this regression line was significantly different from zero, the slope itself was -0.066, suggesting a minimal change from a short tilt to a long tilt duration. This has also been observed in an earlier study(van Campen, Verheugt et al. 2020).

### Limitations

We acknowledge that the present study is retrospective and referral bias by the general practitioner may have played a role, selectively referring patients with orthostatic symptoms. Although this might have affected the generalizability of our CBF measurements to the entire population of individuals with ME/CFS, it would not have interfered with the validity of the observed relationships between CO and CBF. Our study did not enroll those who were bedbound, and we elected not to expose those with more severe functional impairments to tilt testing. Moreover, we only selected those patients with a normal HR and BP response during tilting. The same analysis should be performed in patients with POTS and orthostatic hypotension. Individuals with ME/CFS have been reported to have variable function from day to day and week to week. Future studies can evaluate whether the CBF and CO measurements differ on “good” versus “bad” days. Our focus was on the prevalence of reductions of CBF and CO and therefore mechanisms of CBF and CO changes and regional cerebral blood flow differences were beyond the scope of this study. These topics would be important to investigate further.

Finally, the use of extracranial Doppler flow to measure cerebral blood flow must be replicated by others and in different patient groups. It is unclear how much the orthostatic intolerance of ME/CFS patients differ from other forms of circulatory dysfunction.

## Conclusions

In ME/CFS patients undergoing tilt testing because of the suspicion of orthostatic intolerance, with measurements of both the CO and the CBF, two different patterns were observed: 1) a limited number of patients (9%) had a %CBF reduction in the range of HC. This group of patients had a milder severity of the disease compared to the group of patients with an abnormal %CBF reduction and comprised more males. The largest group of patients (91%) were those with an abnormal %CBF reduction. in the latter group multiple regression analysis with clinical, hemodynamic and PetCO_2_ data showed that the %CO reduction robustly predicted the %CBF reduction, with minor contributions of the PetCO_2_ and the tilt duration. These data suggest that circulatory improvement by increased water and salt intake, compression garments, and medications targeted to improve CO are the primary targets to improve orthostatic intolerance. This needs to be prospectively assessed in randomized, placebo-controlled trials.

## Data Availability

The raw data supporting the conclusions of this article will be made available by the authors, without undue reservation, to any qualified researcher.

## Notes

### Competing Interest Statement

The authors have declared no competing interest.

### Clinical Trial

This is a descriptive study of clinical data, no trial registration was necesaary.

### Funding Statement

This study was performed without grant funding.

### Author Declarations

The study was approved by the medical ethics committee of the Slotervaart Hospital, Amsterdam, for healthy controls P1450 and for ME/CFS patients P1736.

## References

1. Andresen, J., N. I. Shafi and R. M. Bryan, Jr. (2006). “Endothelial influences on cerebrovascular tone.” J Appl Physiol (1985) 100(1): 318–327.

2. Bai, T., S. Yu and J. Feng (2022). “Advances in the Role of Endothelial Cells in Cerebral Small Vessel Disease.” Front Neurol 13: 861714.

3. Bronzwaer, A.-S. G. T., J. Verbree, W. J. Stok, M. J. A. P. Daemen, M. A. van Buchem, M. J. P. van Osch and J. J. van Lieshout (2017). “The cerebrovascular response to lower-body negative pressure vs. head-up tilt.” Journal of Applied Physiology 122(4): 877–883.

4. Brown, C. M., M. Dütsch, M. J. Hecht, B. Neundörfer and M. J. Hilz (2003). “Assessment of cerebrovascular and cardiovascular responses to lower body negative pressure as a test of cerebral autoregulation.” J Neurol Sci 208(1-2): 71–78.

5. Buchwald, D., T. Pearlman, P. Kith and K. Schmaling (1994). “Gender differences in patients with chronic fatigue syndrome.” J Gen Intern Med 9(7): 397–401.

6. Carruthers, B. M., M. I. van de Sande, K. L. DE Meirleir, N. G. Klimas, G. Broderick, T. Mitchell, D. Staines, A. C. Powles, N. Speight, R. Vallings, L. Bateman, B. Baumgarten-Austrheim, D. S. Bell, N. Carlo-Stella, J. Chia, A. Darragh, D. Jo, D. Lewis, A. R. Light, S. Marshall-Gradisbik, I. Mena, J. A. Mikovits, K. Miwa, M. Murovska, M. L. Pall and S. Stevens (2011). “Myalgic encephalomyelitis: International Consensus Criteria.” J Intern Med 270(4): 327–338.

7. Castle-Kirszbaum, M., W. G. Parkin, T. Goldschlager and P. M. Lewis (2022). “Cardiac Output and Cerebral Blood Flow: A Systematic Review of Cardio-Cerebral Coupling.” J Neurosurg Anesthesiol 34(4): 352–363.

8. Claassen, J., D. H. J. Thijssen, R. B. Panerai and F. M. Faraci (2021). “Regulation of cerebral blood flow in humans: physiology and clinical implications of autoregulation.” Physiol Rev 101(4): 1487–1559.

9. Davenport, T. E., M. Lehnen, S. R. Stevens, J. M. VanNess, J. Stevens and C. R. Snell (2019). “Chronotropic Intolerance: An Overlooked Determinant of Symptoms and Activity Limitation in Myalgic Encephalomyelitis/Chronic Fatigue Syndrome?” Front Pediatr 7: 82.

10. Dubchenko, E., A. Ivanov, N. Spirina, N. Smirnova, M. Melnikov, A. Boyko, E. Gusev and A. Kubatiev (2020). “Hyperhomocysteinemia and Endothelial Dysfunction in Multiple Sclerosis.” Brain Sciences 10(9): 637.

11. Eeftinck Schattenkerk, D. W., J. J. van Lieshout, A. H. van den Meiracker, K. R. Wesseling, S. Blanc, W. Wieling, G. A. van Montfrans, J. J. Settels, K. H. Wesseling and B. E. Westerhof (2009). “Nexfin noninvasive continuous blood pressure validated against Riva-Rocci/Korotkoff.” Am J Hypertens 22(4): 378–383.

12. Fang, Y. C., Y. C. Hsieh, C. J. Hu and Y. K. Tu (2023). “Endothelial Dysfunction in Neurodegenerative Diseases.” Int J Mol Sci 24(3).

13. Faro, M., N. Sàez-Francás, J. Castro-Marrero, L. Aliste, T. Fernández de Sevilla and J. Alegre (2016). “Gender differences in chronic fatigue syndrome.” Reumatol Clin 12(2): 72–77.

14. Fedorowski, A., P. Burri and O. Melander (2009). “Orthostatic hypotension in genetically related hypertensive and normotensive individuals.” J Hypertens 27(5): 976–982.

15. Freeman, R., W. Wieling, F. B. Axelrod, D. G. Benditt, E. Benarroch, I. Biaggioni, W. P. Cheshire, T. Chelimsky, P. Cortelli, C. H. Gibbons, D. S. Goldstein, R. Hainsworth, M. J. Hilz, G. Jacob, H. Kaufmann, J. Jordan, L. A. Lipsitz, B. D. Levine, P. A. Low, C. Mathias, S. R. Raj, D. Robertson, P. Sandroni, I. J. Schatz, R. Schondorf, J. M. Stewart and J. G. van Dijk (2011). “Consensus statement on the definition of orthostatic hypotension, neurally mediated syncope and the postural tachycardia syndrome.” Auton Neurosci 161(1-2): 46–48.

16. Fu, Q., T. B. Vangundy, M. M. Galbreath, S. Shibata, M. Jain, J. L. Hastings, P. S. Bhella and B. D. Levine (2010). “Cardiac origins of the postural orthostatic tachycardia syndrome.” J Am Coll Cardiol 55(25): 2858–2868.

17. Fukuda, K., S. E. Straus, I. Hickie, M. C. Sharpe, J. G. Dobbins and A. Komaroff (1994). “The chronic fatigue syndrome: a comprehensive approach to its definition and study. International Chronic Fatigue Syndrome Study Group.” Ann Intern Med 121(12): 953–959.

18. Hoiland, R. L., J. A. Fisher and P. N. Ainslie (2019). “Regulation of the Cerebral Circulation by Arterial Carbon Dioxide.” Compr Physiol 9(3): 1101–1154.

19. Institute Of Medicine (IOM), Ed. (2015). Beyond mayalgic encephalomyelitis/chronic fatigue syndrome: redefining an illness. Washington DC, The National Academies Press.

20. Keskin, K., S. Çiftçi, J. Öncü, G. Melike Doğan, G. Çetinkal, S. Sezai Yıldız, S. Sığırcı and K. Orta Kılıçkesmez (2021). “Orthostatic hypotension and age-related sarcopenia.” Turk J Phys Med Rehabil 67(1): 25–31.

21. Kollár, B., A. Blaho, K. Valovičová, M. Poddaný, P. Valkovič, I. Straka, P. Turčáni and P. Šiarnik (2022). “Impairment of endothelial function in Parkinson’s disease.” BMC Research Notes 15(1): 284.

22. Laerd Statistics (2015) “Leard Statistics. Multiple regression using SPSS Statistics. Statistical tutorials and software guides. Retrieved from https://statistics.laerd.com/.”

23. Maes, F., S. Pierard, C. de Meester, J. Boulif, M. Amzulescu, D. Vancraeynest, A.-C. Pouleur, A. Pasquet, B. Gerber and J.-L. Vanoverschelde (2017). “Impact of left ventricular outflow tract ellipticity on the grading of aortic stenosis in patients with normal ejection fraction.” Journal of Cardiovascular Magnetic Resonance 19(1): 37.

24. Martina, J. R., B. E. Westerhof, J. van Goudoever, E. M. de Beaumont, J. Truijen, Y. S. Kim, R. V. Immink, D. A. Jobsis, M. W. Hollmann, J. R. Lahpor, B. A. de Mol and J. J. van Lieshout (2012). “Noninvasive continuous arterial blood pressure monitoring with Nexfin(R).” Anesthesiology 116(5): 1092–1103.

25. McLaughlin, M., N. E. M. Sanal-Hayes, L. D. Hayes, E. C. Berry and N. F. Sculthorpe (2023). “People with Long COVID and Myalgic Encephalomyelitis/Chronic Fatigue Syndrome Exhibit Similarly Impaired Vascular Function.” Am J Med.

26. Meng, L., W. Hou, J. Chui, R. Han and A. W. Gelb (2015). “Cardiac Output and Cerebral Blood Flow: The Integrated Regulation of Brain Perfusion in Adult Humans.” Anesthesiology 123(5): 1198–1208.

27. Nakatomi, Y., K. Mizuno, A. Ishii, Y. Wada, M. Tanaka, S. Tazawa, K. Onoe, S. Fukuda, J. Kawabe, K. Takahashi, Y. Kataoka, S. Shiomi, K. Yamaguti, M. Inaba, H. Kuratsune and Y. Watanabe (2014). “Neuroinflammation in Patients with Chronic Fatigue Syndrome/Myalgic Encephalomyelitis: An 11C- (R)-PK11195 PET Study.” J Nucl Med 55(6): 945–950.

28. Newton, D. J., G. Kennedy, K. K. Chan, C. C. Lang, J. J. Belch and F. Khan (2012). “Large and small artery endothelial dysfunction in chronic fatigue syndrome.” Int J Cardiol 154(3): 335–336.

29. Nunes, J. M., A. Kruger, A. Proal, D. B. Kell and E. Pretorius (2022). “The Occurrence of Hyperactivated Platelets and Fibrinaloid Microclots in Myalgic Encephalomyelitis/Chronic Fatigue Syndrome(ME/CFS).” Pharmaceuticals (Basel) 15(8).

30. Phillips, A. A., F. H. Chan, M. M. Zheng, A. V. Krassioukov and P. N. Ainslie (2016). “Neurovascular coupling in humans: Physiology, methodological advances and clinical implications.” J Cereb Blood Flow Metab 36(4): 647–664.

31. Rowe, P. C., D. F. Barron, H. Calkins, I. H. Maumenee, P. Y. Tong and M. T. Geraghty (1999). “Orthostatic intolerance and chronic fatigue syndrome associated with Ehlers-Danlos syndrome.” J Pediatr 135(4): 494–499.

32. Sandvik, M. K., K. Sørland, E. Leirgul, I. G. Rekeland, C. S. Stavland, O. Mella and Ø. Fluge (2023). “Endothelial dysfunction in ME/CFS patients.” PLoS One 18(2): e0280942.

33. Scherbakov, N., M. Szklarski, J. Hartwig, F. Sotzny, S. Lorenz, A. Meyer, P. Grabowski, W. Doehner and C. Scheibenbogen (2020). “Peripheral endothelial dysfunction in myalgic encephalomyelitis/chronic fatigue syndrome.” ESC Heart Failure 7(3): 1064–1071.

34. Sheldon, R. S., B. P. Grubb, 2nd, B. Olshansky, W. K. Shen, H. Calkins, M. Brignole, S. R. Raj, A. D. Krahn, C. A. Morillo, J. M. Stewart, R. Sutton, P. Sandroni, K. J. Friday, D. T. Hachul, M. I. Cohen, D. H. Lau, K. A. Mayuga, J. P. Moak, R. K. Sandhu and K. Kanjwal (2015). “2015 heart rhythm society expert consensus statement on the diagnosis and treatment of postural tachycardia syndrome, inappropriate sinus tachycardia, and vasovagal syncope.” Heart Rhythm 12(6): e41–63.

35. Silverman, A. and N. H. Petersen (2024). Physiology, Cerebral Autoregulation. StatPearls. Treasure Island (FL), StatPearls Publishing

36. Copyright © 2024, StatPearls Publishing LLC.

37. Sørland, K., M. K. Sandvik, I. G. Rekeland, L. Ribu, M. C. Småstuen, O. Mella and Ø. Fluge (2021). “Reduced Endothelial Function in Myalgic Encephalomyelitis/Chronic Fatigue Syndrome–Results From Open-Label Cyclophosphamide Intervention Study.” Frontiers in Medicine 8.

38. Stewart, J. M., P. Pianosi, M. A. Shaban, C. Terilli, M. Svistunova, P. Visintainer and M. S. Medow (2018). “Postural Hyperventilation as a Cause of Postural Tachycardia Syndrome: Increased Systemic Vascular Resistance and Decreased Cardiac Output When Upright in All Postural Tachycardia Syndrome Variants.” J Am Heart Assoc 7(13): e008854.

39. Streeten, D. H., D. Thomas and D. S. Bell (2000). “The roles of orthostatic hypotension, orthostatic tachycardia, and subnormal erythrocyte volume in the pathogenesis of the chronic fatigue syndrome.” Am J Med Sci 320(1): 1–8.

40. Timmers, H. J., W. Wieling, P. M. Soetekouw, G. Bleijenberg, J. W. Van Der Meer and J. W. Lenders (2002). “Hemodynamic and neurohumoral responses to head-up tilt in patients with chronic fatigue syndrome.” Clin Auton Res 12(4): 273–280.

41. Tseng, C.-L. and B. H. Natelson (2004). “Few Gender Differences Exist Between Women and Men with Chronic Fatigue Syndrome.” Journal of Clinical Psychology in Medical Settings 11(1): 55–62.

42. van Campen, C., P. C. Rowe, F. W. A. Verheugt and F. C. Visser (2023). “Influence of end-tidal CO(2) on cerebral blood flow during orthostatic stress in controls and adults with myalgic encephalomyelitis/chronic fatigue syndrome.” Physiol Rep 11(17): e15639.

43. van Campen, C., P. C. Rowe and F. C. Visser (2023). “Worsening Symptoms Is Associated with Larger Cerebral Blood Flow Abnormalities during Tilt-Testing in Myalgic Encephalomyelitis/Chronic Fatigue Syndrome (ME/CFS).” Medicina (Kaunas) 59(12).

44. van Campen, C., F. W. A. Verheugt, P. C. Rowe and F. C. Visser (2023). “Orthostatic chronotropic incompetence in patients with myalgic encephalomyelitis/chronic fatigue syndrome (ME/CFS).” IBRO Neurosci Rep 15: 1–10.

45. van Campen, C. L. M. C., P. C. Rowe and V. F.C. (2023). “Comparison of a 20 degree and 70 degree tilt test in adolescent myalgic encephalomyelits/chronic fatigue syndrome (ME/CFS) patients.” Frontiers in Pediatrics 11: 1–8.

46. van Campen, C. L. M. C., P. C. Rowe and F. C. Visser (2018). “Blood Volume Status in ME/CFS Correlates With the Presence or Absence of Orthostatic Symptoms: Preliminary Results.” Front Pediatr 6: 4.

47. van Campen, C. L. M. C., P. C. Rowe and F. C. Visser (2020). “Cerebral Blood Flow Is Reduced in Severe Myalgic Encephalomyelitis/Chronic Fatigue Syndrome Patients During Mild Orthostatic Stress Testing: An Exploratory Study at 20 Degrees of Head-Up Tilt Testing.” Healthcare (Basel) 8(2): 169.

48. van Campen, C. L. M. C., P. C. Rowe and F. C. Visser (2020). “Reductions in Cerebral Blood Flow Can Be Provoked by Sitting in Severe Myalgic Encephalomyelitis/Chronic Fatigue Syndrome Patients.” Healthcare 8: 394.

49. van Campen, C. L. M. C., P. C. Rowe and F. C. Visser (2021). “Cerebral blood flow remains reduced after tilt testing in myalgic encephalomyelitis/chronic fatigue syndrome patients.” Clinical Neurophysiology Practice 6: 245–255.

50. van Campen, C. L. M. C., P. C. Rowe and F. C. Visser (2021). “Compression Stockings Improve Cardiac Output and Cerebral Blood Flow during Tilt Testing in Myalgic Encephalomyelitis/Chronic Fatigue Syndrome (ME/CFS) Patients: A Randomized Crossover Trial.” Medicina (Kaunas) 58(1).

51. van Campen, C. L. M. C., P. C. Rowe and F. C. Visser (2021). “The Myalgic Encephalomyelitis/Chronic Fatigue Syndrome Patients with Joint Hypermobility Show Larger Cerebral Blood Flow Reductions during Orthostatic Stress Testing Than Patients without Hypermobility: A Case Control Study.” Medical Research Archives; Vol 9 No 6 (2021): Vol.9 Issue 6, June,2021DO - 10.18103/mra.v9i6.2494.

52. van Campen, C. L. M. C., P. C. Rowe and F. C. Visser (2021). “Orthostatic Symptoms and Reductions in Cerebral Blood Flow in Long-Haul COVID-19 Patients: Similarities with Myalgic Encephalomyelitis/Chronic Fatigue Syndrome.” Medicina (Kaunas) 58(1).

53. van Campen, C. L. M. C., F. W. A. Verheugt, P. C. Rowe and F. C. Visser (2020). “Cerebral blood flow is reduced in ME/CFS during head-up tilt testing even in the absence of hypotension or tachycardia: A quantitative, controlled study using Doppler echography.” Clin Neurophysiol Pract 5: 50–58.

54. van Campen, C. L. M. C., F. W. A. Verheugt, P. C. Rowe and F. C. Visser (2021). “Comparison of the finger plethysmography derived stroke volumes by Nexfin CO Trek and suprasternal aortic Doppler derived stroke volume measruements in adults with myalgic encephaolomyelitis/chronic fatgitue syndrome and in healthy controls.” Technology and Health Care 29(4): 629–642.

55. van Campen, C. L. M. C., F. W. A. Verheugt and F. C. Visser (2018). “Cerebral blood flow changes during tilt table testing in healthy volunteers, as assessed by Doppler imaging of the carotid and vertebral arteries.” Clin Neurophysiol Pract 3: 91–95.

56. van Campen, C. L. M. C., F. W. A. Verheugt and F. C. Visser (2018). “Quantification of the beneficial effects of compression stockings on symptoms of exercise and orthostatic intolerance in chronic fatigue/myalgic encephalomyelitis patients.” International Journal of Clinical Medicine 9: 367–376.

57. van Campen, C. L. M. C. and F. C. Visser (2018). “The abnormal Cardiac Index and Stroke Volume Index changes during a normal Tilt Table Test in ME/CFS patients compared to healthy volunteers, are not related to deconditioning.” Journal Of Thrombosis and Circulation (2): 1–8.

58. van Campen, C. L. M. C., F. C. Visser, C. C. de Cock, H. S. Vos, O. Kamp and C. A. Visser (2006). “Comparison of the haemodynamics of different pacing sites in patients undergoing resynchronisation treatment: need for individualisation of lead localisation.” Heart 92(12): 1795–1800.

59. Wang, X. L., T. Y. Ling, M. C. Charlesworth, J. J. Figueroa, P. Low, W. K. Shen and H. C. Lee (2013). “Autoimmunoreactive IgGs against cardiac lipid raft-associated proteins in patients with postural orthostatic tachycardia syndrome.” Transl Res 162(1): 34–44.

60. Willie, C. K., Y. C. Tzeng, J. A. Fisher and P. N. Ainslie (2014). “Integrative regulation of human brain blood flow.” J Physiol 592(5): 841–859.

61. Wirth, K. J., C. Scheibenbogen and F. Paul (2021). “An attempt to explain the neurological symptoms of Myalgic Encephalomyelitis/Chronic Fatigue Syndrome.” Journal of Translational Medicine 19(1): 471.

62. Wyller, V. B., V. Vitelli, D. Sulheim, E. Fagermoen, A. Winger, K. Godang and J. Bollerslev (2016). “Altered neuroendocrine control and association to clinical symptoms in adolescent chronic fatigue syndrome: a cross-sectional study.” J Transl Med 14(1): 121.

